# Transition in learning approach for undergraduate medical students of Bangladesh in Covid 19 pandemic: A situation analysis

**DOI:** 10.1101/2021.04.29.21256358

**Authors:** Fatiha Tasmin Jeenia, Md Jamal Uddin Tanin, Jannatul Ferdoush, Fatema Johora, Afroza Hoque, Asma Akter Abbasy, Halima Sadia, Rehnuma Urmi, Priyanka Moitra, Quazi Saheli Sarah, Maliha Ata, Kohinoor Parveen

## Abstract

**Background:** The outbreak of Covid-19 pandemic has fundamentally transformed the landscape of medical education system upside down worldwide. This unanticipated transition without any pre build infrastructure has made this altered prospect more challenging in Bangladesh. Though many countries across the world utilize Web Based Learning (WBL), but medical students of Bangladesh are mostly unfamiliar and unaccustomed with this newly imposed online learning avenue. Therefore, this study has evaluated the familiarity, usage, attitude of students towards online class and figured out the barriers witnessed by students in Bangladesh prospect.

**Methods:** This cross sectional, questionnaire based study was conducted in medical colleges across Bangladesh. A questionnaire linked to google form were distributed to undergraduate medical students all over Bangladesh through different social platforms. The answered questionnaires were automatically stored in Google drive in a specific email ID.

**Results:** A total of 1708 students participated. Among the respondents 45.1% were satisfied with online class. Though most students (45.8%) think online class is not effective like traditional lectures but 47.4% agrees to the point that online class should have complementary role in medical education. One of the strong attitude of medical students revealed that, most of them undoubtedly in unison (49.5% disagree, 30.3% strongly disagree) with that online class can never replace traditional class. 77.2% students responded that web based learning is interactive. 54.9% students pointed out interrupted internet connections with low speed during class which is a barrier to WBL. 83.2% of the respondents complained about facing audio visual problem in class which is attributed to poor network connectivity. Most students (74.8%) found online classes costly and 53.8% needed technical supports for online class.

**Conclusion:** This study finding can suggest a potential reform for online class in Bangladesh. Addressing the obstacles and expectations can execute a fruitful web based learning in Bangladesh.

## Introduction

Unanticipated transition of medical education system from traditional to a complete web based approach is quite challenging. And the outbreak of Covid 19 pandemic has transformed medical education system in this direction worldwide. While the pre-clinical classes are completely switched to electronic learning, the clinical students are largely propelled voluntarily to join front liners in countries like UK, USA and others. And this mobilization is considered as to provide a surplus of unique opportunity to be a self-directed learner and developing overall clinical skills regarding pandemic management^[1,2]^. Though the online teaching learning is ongoing currently in a rigorous manner but ineffective online learning strategies, poor motivation of students, suboptimal communication skills, students from low socioeconomic strata with lack of equipment and connectivity has brought additional challenges. Besides, in this online learning platform a two way teacher student interaction is quite difficult to replicate like the real time forum^[3,4]^. In order to overcome the challenges of this looming new normal web based learning worldwide, many medical educators are ruminating meticulously on how best to ensure an uninterrupted and stringent medical education and clinical training that will produce a steady stream of future competent physician^[5,6]^. Technology is evolving over years along with modern revolution. The learning system needs to be tailored accordingly. Nonetheless, learning approach has transformed towards internet based or web based system in last few decades. ‘Web based learning’ often called online learning or e-learning as it includes online course content. “Electronic (e) or online learning can be defined as the utilization of electronic technology and media to deliver, support and enhance both teaching and learning and it involves communication between learners and teachers using online content”^[7,8]^. Live lectures (video streaming), videoconferencing, webinars, discussion forums via email are all possible through the web^[9]^. Contents can be delivered in the form of pure distance learning, blended with face to face learning, virtual classroom, satellite television, audio or video tape^[10]^. Web-based learning now a days is a popular approach worldwide. Advantages of web based learning includes its ability to link resources in many different formats^[11]^, Can be an efficient way of delivering course materials, resources can be made available from any location and at any time, has potential for widening access for example, to part time, or work based students, can encourage more independent and active learning, can provide a useful source of supplementary materials for conventional programs. Aside than that, web based learning has several drawbacks. Inadequate access to appropriate computer equipment can be a problem for students. Learners sometimes find it frustrating if they cannot access images, graphics and video clips because of poor equipment. The necessary infrastructure must be available and affordable in this regard. Information can vary in quality and accuracy, therefore, guidance and signposting is needed. Besides, students might feel isolated^[12]^. Particularly, medical education system is mostly practical oriented as it involves clinical settings. Therefore, a sole online learning is not convenient in medical education. There are many contradictions regarding the compatibility and acceptability of web based learning in medical education. A handful of researches has been conducted both in undergraduate and postgraduate medical classes with regard to determine the versatility of web based learning. Two school of thoughts on this ground persists. Majority of the research revealed the students complacency with the online education and advocacy to blend web learning with traditional one. The acceptance are mostly observed in European and Western countries in a blended form with traditional approach. Developed countries like England, Germany, and the United States already incorporated information technology into educational settings to support teaching and learning since the 1980s^[13]^. In developing countries such like Taiwan, e-learning has gained public awareness since 2000 through the popularity of educational resources websites. Asian countries particularly South East Asian countries lagging behind in this aspect. Few researches revealed the usage of web based learning to educate rural population in South Asian countries like India, Sri Lanka and Bhutan^[14]^. Research found out the potential of virtual classes in Bangladesh^[15]^. However, In Bangladesh web based learning in medical education got a pace in this Covid 19 pandemic which is giving the golden opportunity to incorporate information technology more precisely into medical education. One of the prerequisite is to explore the scope and dilemmas of WBL in prospect of Bangladesh in order to overcome the shortcomings. In this backdrop, this study will analyze the situation of online learning, find out the student’s attitude, response to this transition from traditional to web based learning and will try to figure out the obstacles of web based learning.

## Materials and Methods

### Study Sites and Participants

This study was conducted on all phase (Phase 1 to phase 4) undergraduate medical students of all government and non-government medical colleges of Bangladesh from August 2020 to January 2020. Voluntarily willing students were the participants of this study. Students who avoided the questionnaire and were unwilling to fill up did not participate in this study. The study was approved by Institutional Review Board in compliance with the provision of Declaration of Helsinki.

### Study Design

This study was a cross sectional type of observational study.

### Study Procedure

A questionnaire was developed and was linked in google form. In the questionnaire, questions were arranged under three headings to assess the scenario of undergraduate online medical class that first ever occurred in Bangladesh in a large scale. The questionnaire was distributed among students of Government and Non-Government Medical Colleges of Bangladesh through email, Messenger app, WhatsApp and other social platforms to access students all around Bangladesh. Those students who were interested to the questionnaire participated in the study and submitted their response. In order to avoid duplicated response from a single participant, the automated google form questionnaire accepted only one response from a single mail account. A reminder mail or message was sent on 7^th^ day and 15^th^ day of the primary one. The response generated by the students was received through google drive.

### Outcome Measures

The primary outcome measure of the study was to figure out the attitude of students towards web based learning. The secondary outcome measures were identifying the percentage of students accustomed to WBL, exploring the percentage of students satisfied with online classes, determining students percentage accustomed to web based learning, sorting out percentage of online platform, electronic gadget and network connectivity used by the students for web based learning, identifying the problems and barriers faced by students while attending online classes.

### Statistical analysis

All the answered question were transferred from google drive into a spreadsheet. Frequency distribution and percentage was calculated by using SPSS version 22.

## Results and Observations

A total of 1708 students participated in this study. Among them, 622 (36.4%) students were from phase 1, 400 (23.4%) from phase 2, 471 (27.6%) and 214 (12.5%) from phase 3 and phase 4 respectively (Figure 1).

**Figure 1:**
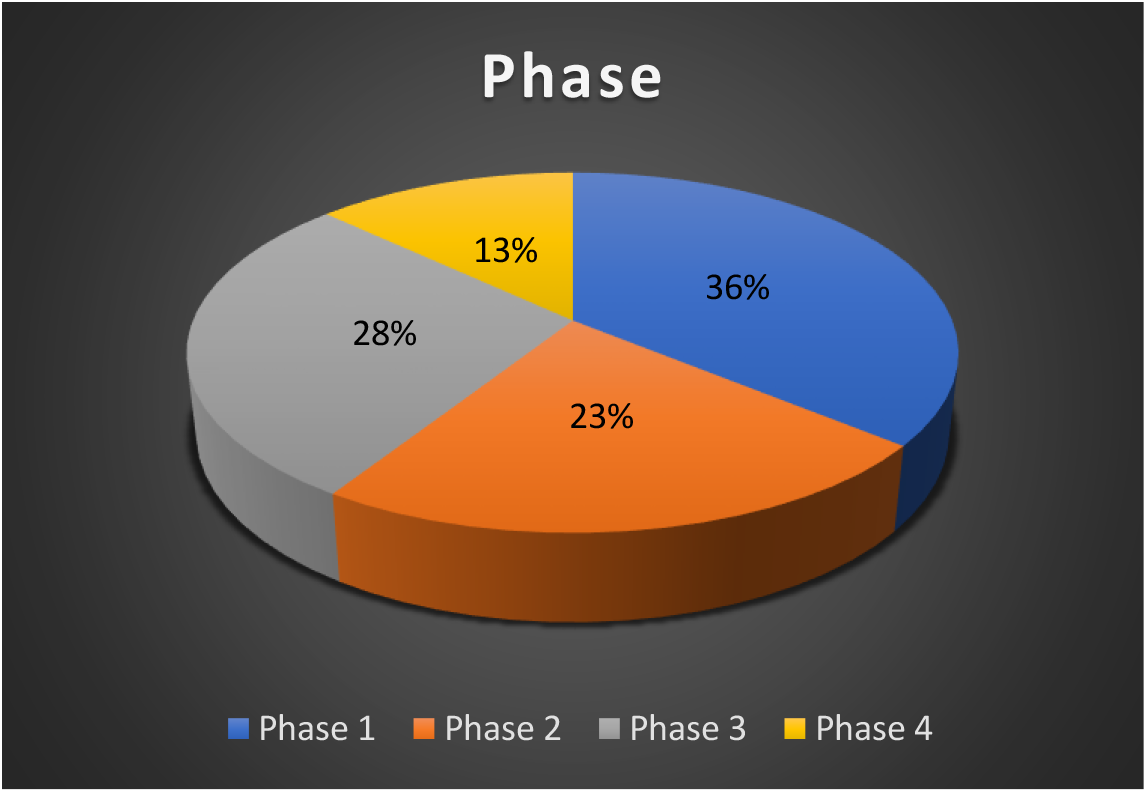
Distribution of undergraduate medical students according to phase.

### Familiarity and usage of web based learning

Figure 2 is showing, 67% students are familiar with web based learning and only 37% students attended online class prior to pandemic particularly arranged non-institutionally and in the form of scattered topics (Figure 3).

**Figure 2:**
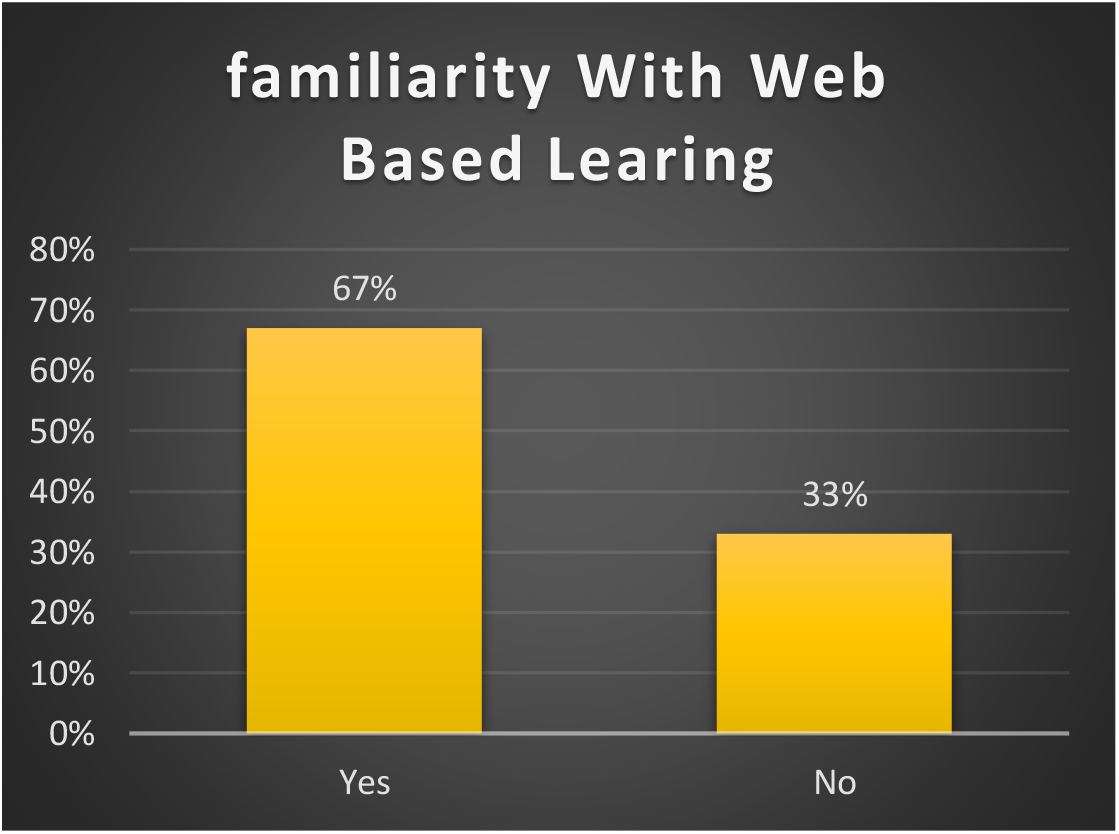
Percentage of students familiar with web based learning.

**Figure 3:**
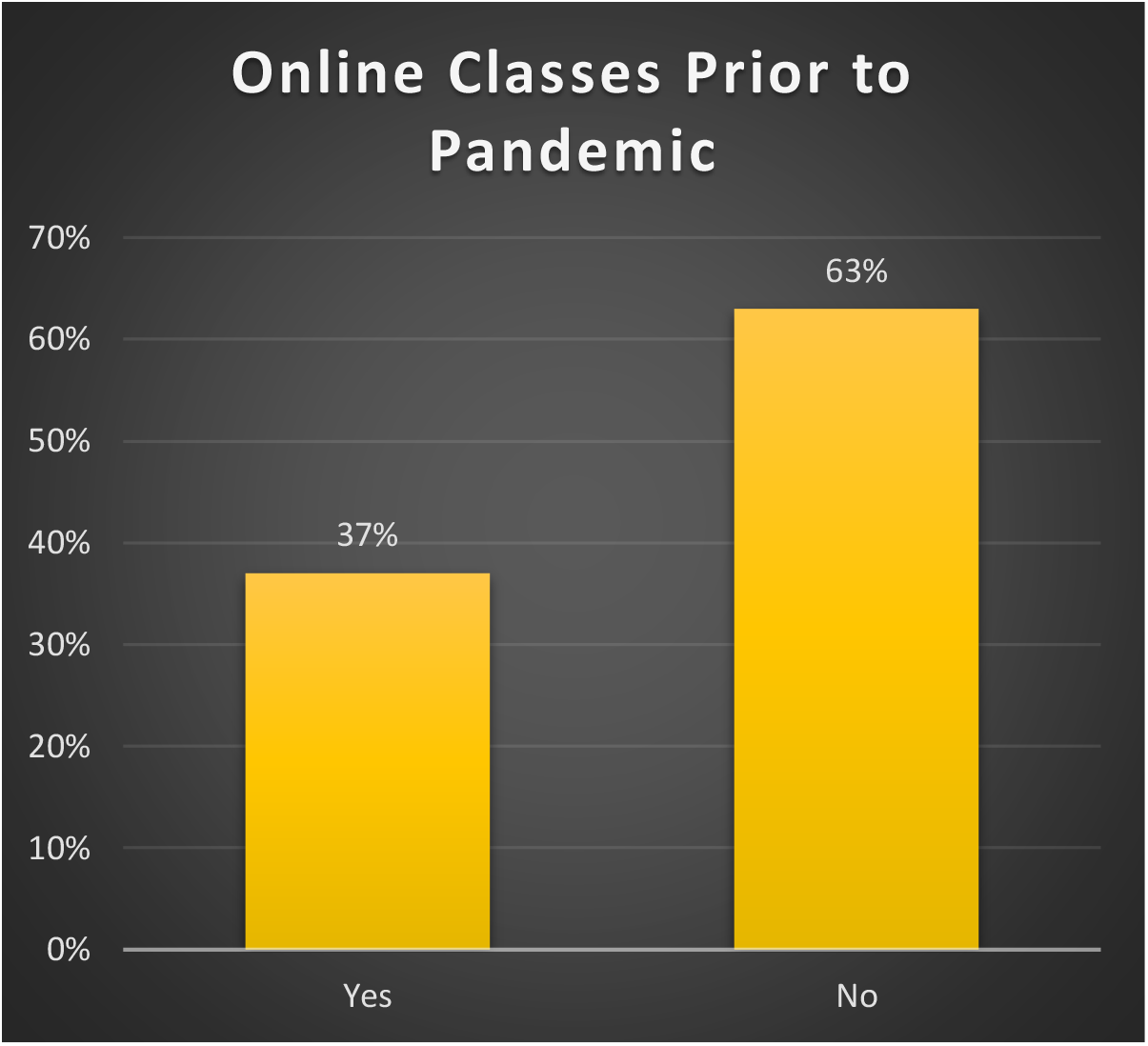
Percentage of students attended online classes prior to pandemic.

Most of the students use smart phone to attend online class. The frequency of gadget used by students are shown in Table 1.

**Table 1:** Types of gadgets used by the students for online class.

| Gadget | Frequency | Percentage |
| --- | --- | --- |
| Smart phone | 1500 | 87.8 |
| Laptop | 200 | 11.7 |
| Desktop | 08 | 0.5 |

53% of the students uses mobile data package for internet connection and 47% used broadband connection (Figure 4).

**Figure 4:**
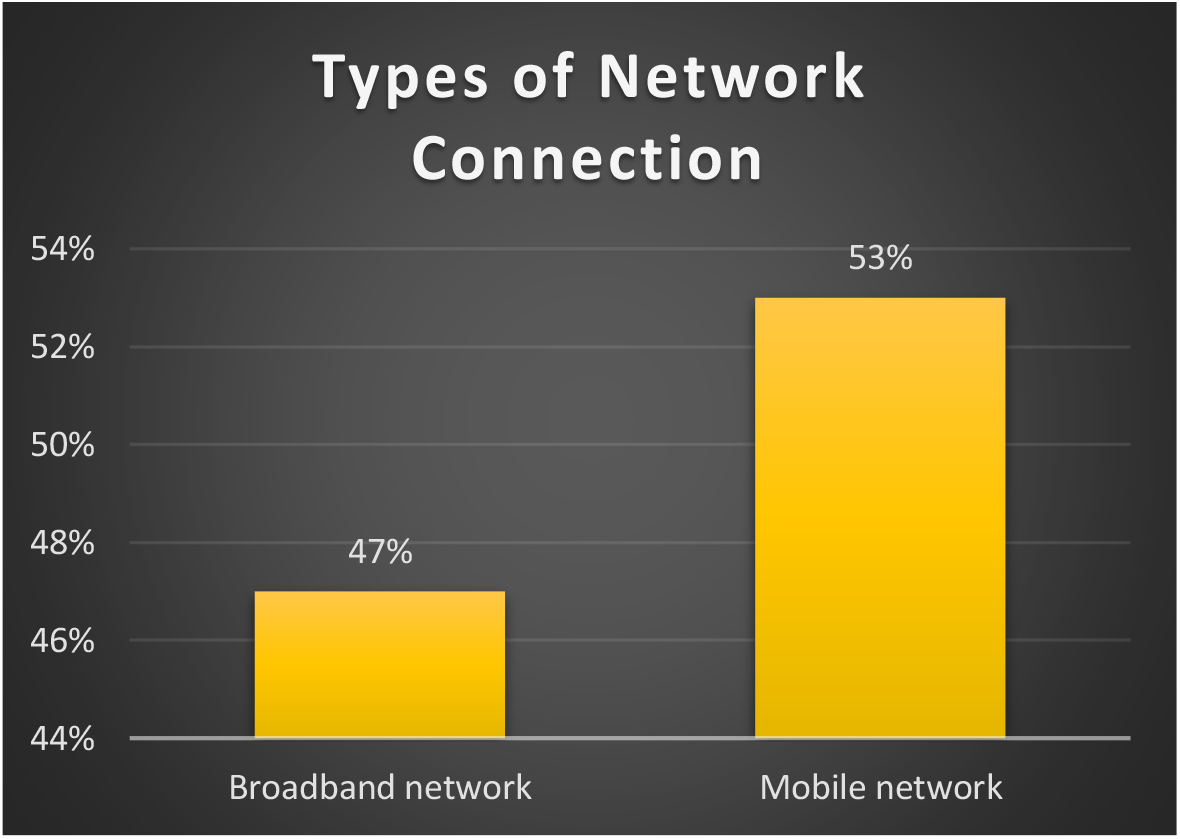
Types of network connectivity used by students for online class.

Most of the students prefer online class lecture deliberation to be in the form of power point presentation (67%) followed by lecture using white board and others (15%) (Figure 5).

**Figure 5:**
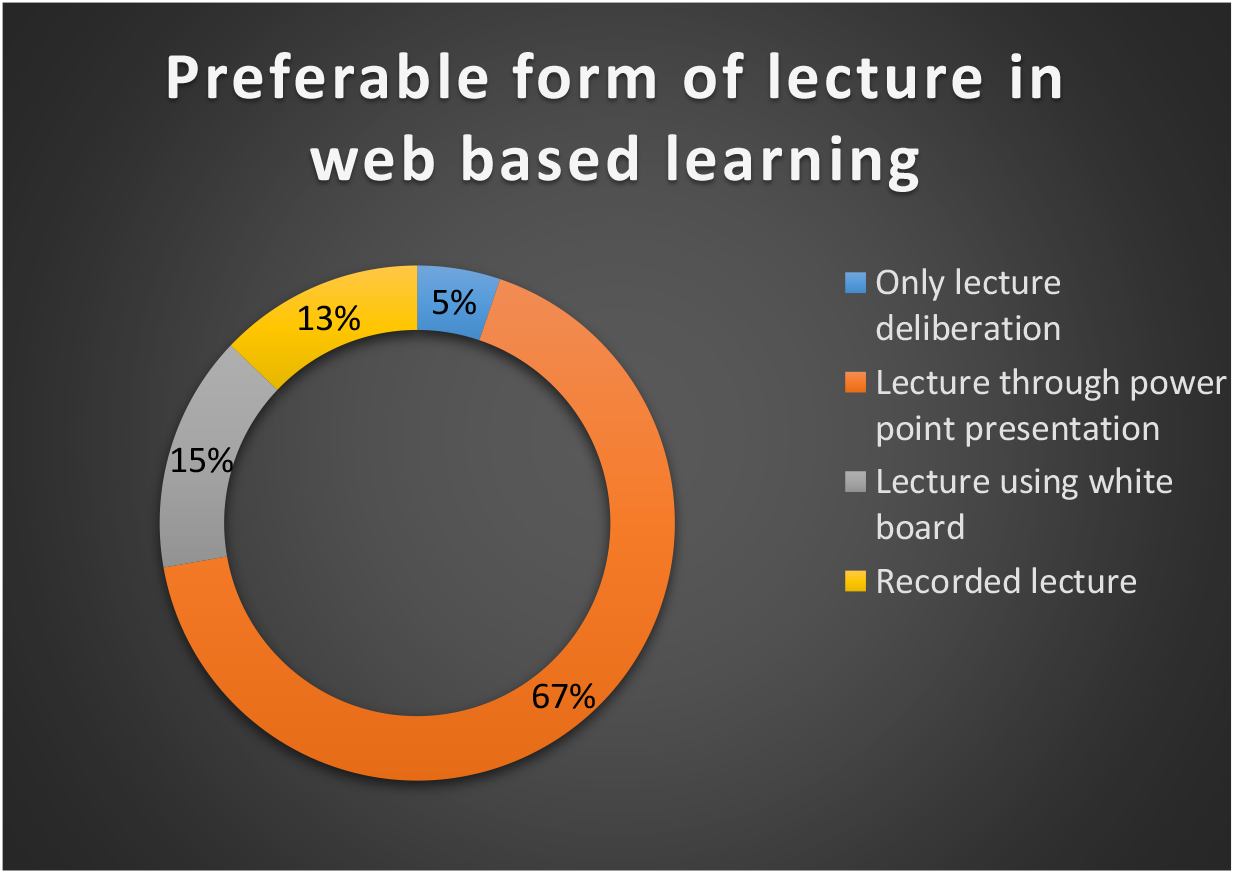
Lecture form preference by students.

Attitudes of medical students regarding web based learning in pandemic were speculated by answering some questions which are shown in Table 2.

**Table 2:** Attitude of students towards web based learning:

| **Questions** | **Response** |  |  |  |
| --- | --- | --- | --- | --- |
|  | **Strongly agree** | **Agree** | **Disagree** | **Strongly disagree** |
| I am satisfied with online classes | 74<br>(4.3%) | 771<br>(45.1%) | 620<br>(36.3%) | 242<br>(14.2%) |
| Web based learning is effective for medical education | 64<br>(3.7%) | 595<br>(34.8%) | 782<br>(45.8%) | 266<br>(15.6%) |
| Web based learning should have complementary role in medical education | 73<br>(4.3%) | 810<br>(47.4%) | 627<br>(36.7%) | 197<br>(11.5%) |
| Web based learning is useful in medical education | 55<br>(3.2%) | 610<br>(35.7%) | 701<br>(41%) | 341<br>(20%) |
| Web based learning can replace the traditional lecture | 42<br>(2.5%) | 302<br>(17.7%) | 846<br>(49.5%) | 517<br>(30.3%) |
| Web based learning must be more prevalent in medical education | 98<br>(5.7%) | 705<br>(41.3%) | 697<br>(40.8%) | 207<br>(12.1%) |
| Web based learning is interactive | 41<br>(2.4%) | 1319<br>(77.2%) | 311<br>(18.2%) | 37<br>(2.2%) |

Barriers of online classes that are identified by the students are shown in Table 3.

**Table 3:** Barriers in web based learning:

| Questions | Response |  |
| --- | --- | --- |
|  | Yes | No |
| Do you have to go to institution or away from home to attend online class in pandemic? | 448 (26.2%) | 1259 (73.7%) |
| Do you get uninterrupted internet access during class time? | 770 (45.1%) | 937 (54.9%) |
| Is there audio visual problems during online class? | 1421 (83.2%) | 286 (16.7%) |
| Do you find online classes costly? | 1277 (74.8%) | 430 (25.2%) |
| Do you think time duration is sufficient for a single class? | 1264 (74%) | 443 (25.9%) |
| Can you ask questions in the class? | 1359 (79.6%) | 348 (20.4%) |
| Do you get pdf of online class? | 815 (47.7%) | 892 (52.2%) |
| Do you need technical support for online class? | 919 (53.8%) | 788 (46.1%) |

## Discussion

In a world infirmed with covid 19 pandemic, the dynamicity of medical education almost slackened in term of practical and clinical orientation which is a mandate for this education system. There is a huge shift of medical education towards online learning all round the world. In Bangladesh where acquiescence of web based learning was very negligible in undergraduate medical education, a sudden notable transition has been observed in this pandemic. This study tried to look into the situation along with student’s perception, attitude and dilemmas in this newly imposed way of education.

This study found that, 98.1% among the responders has attended online class in this pandemic. Presumably, this higher percentage of online class participation is due to having a smart phone in possession of students. 87.8% of the respondents of this study possess a smart phone where as 11.6% and 0.4% have laptop and desktop respectively. 67% students are familiar with WBL as they are attending online classes. 63% students never participated an online class before this pandemic. Precisely, medical colleges does not have provisions for any online classes for undergraduate students in Bangladesh. However, several online organizations occasionally offers short term online courses or classes on a single topic to students. In pandemic most of the educational institutions transformed their academic activities towards web based learning. A similar study of Nepal revealed 98.1% of the students have attended online classes in pandemic and this study also compared the large number of students having smart phone (74.6%) laptop (48.8%) and desktop (1%) as an influencing factor to the higher involvement in online class^[16]^.

Majority of the students (53%) use mobile network for internet connectivity during online class. On the other hand, 47% take on broadband network. Bulk portion of the students (83%) attended online class using zoom as online platform and prefers lecture deliberation by power point presentation (67%) for online class. A study also reported zoom as the most commonly used single platform for education in pandemic^[17]^. 62.8% students used 4G mobile network and 25% used broadband^[18]^.

45.1% of the students are satisfied with the online classes and 36.3% are dissatisfied. Though most of the students (45.8%) think online class in medical education is not effective like traditional lectures but many of them (47.4%) agree to the point that online classes should have complementary role in medical education. One of the strong attitude of medical students regarding web based learning revealed in this study that, most of them undoubtedly in unison (49.5% disagree, 30.3% strongly disagree) with that web based learning can never replace traditional lecture class in medical education. Literally, medical education set up is a more practical and clinical oriented rather solely lecture based system. And students respond in a more interactive way while physically in the class with teachers as well with classmates. Other than that, web based learning bestows the opportunity to attend a class in a suitable time at homely environment. Besides, WBL also brings up the prospect to learn a wide sort of lectures on a single topic from mentors around the country which is not possible with traditional learning. These might be the reason of students’ positive attitude towards WBL to play complementary role in medical education even though more than half of them are dissatisfied with online class.

Ghanizadeh et al., 2018 has found a result which is in concordance with this study regarding the student’s agreement (57.9%) with the more prevalence of e learning in medical education. And students in that study also responded (39.5% agree, 35.6% strongly agree) with the need of e learning to play complementary role in medical education. Besides, Iranian medical students (58.3%) agree that e learning can never replace traditional lecture class and this particular finding coincides with the current study^[19]^. Though medical students in Saudi Arabia think different in this regard from current study. They responded with the better acceptance of online class than campus based class as they consider the recorded lecture very helpful^[20]^. In a study, Polish students found online class less effective than face to face class in terms of increasing skills and social competencies. And they (70%) also renders e learning is less interactive^[21]^. Similarly, in a single center study of Nepal, students referred online class as not effective at all^[16]^.

Meanwhile, in Libya, 24% students are satisfied with the e learning contents whereas 47% students disagree with the satisfaction of e learning content. And another finding consistent with this study is that, most of Libyan students (53%) disagrees with the possible substitution of standard medical education with e learning and only 27% agrees with that ^[18]^. Correspondingly, 26.8% students in Jordan are satisfied with online class ongoing in the midst of pandemic but most of them (44.4%) are not satisfied at all with that^[17]^. A large scale systematic review including 59 studies and over 6000 undergraduate medical students was carried out in 2014 revealed no difference in satisfaction with online learning over traditional approach ^[22]^.

When it comes to the question whether students can interact with teacher in online class, students mostly (77.2%) responded in agreement in this study. 18.2% do not think online class as a good platform to interact with teachers. 42% of the Libyan students also think alike to this study in this particular opinion and 26% considers online class as not interactive ^[18]^.

Current study tried to look into the barriers of web-based learning in Bangladesh reported by the students. As primarily there was nationwide lockdown due to pandemic at the beginning most students (73.8%) could attend class from home. But 26.3% reported that they had to go away from home in order to participate online class. Perhaps, this portion of students resides in rural area where internet connection is very poor and they had to shift place to get a good network. Alongside, 54.9% students pointed out that they experienced interrupted internet connections with low internet speed during class time which is a barrier to WBL. 45.1% said they got uninterrupted internet access in online class time. In a study of UK on 2721 students, they assigned ‘no travel’ as an advantage of WBL. Though 21.53% of them identified poor internet connection as a barrier to attend online classes ^[23]^. In Nepal, some students (31.6%) denoted electricity cut down as the factor behind internet disturbance, 47.4% and 19.6% found satisfactory and good internet connection respectively ^[16]^.

83.2% of the respondents complained about facing audio visual problem during online class and this is attributed to the poor network connectivity and interrupted internet service throughout the class time. Most students in Bangladesh (74.8%) found online classes costly and this particular finding owing to the accessibility of 4G mobile internet data packages which are quite expensive for the students. Another barrier need to be mentioned is 53.8% of the students needed technical supports for continued online class. Precisely, most of the institutions do not have infrastructure for supporting WBL and no provision of training facilities or technological support for the students as well.

An Indian study on 1068 students observed poor internet accessibility with audio visual problem, costly internet package ascribed to increased cost for online class as barriers ^[24]^ and this observation actually supports this study. Another study conducted in Sri Lanka also pointed lack of reliable network infrastructure as one of the major constraint for WBL ^[25]^. An integrative review comprised over 3000 studies identified poor technical skills, inadequate infrastructure including poor internet access, high cost internet connection and packages not favorable for students, absence of institutional strategies and support for providing technical supports to students as obstacles of online learning in medical education and all this findings are in concordance with the current study^[26]^.

## Conclusion

With increasing severe Covid cases Bangladesh is now going through a second wave and medical education is still following web based system after several times declaration from government for strict closure of educational institutions. It seems the stagnant condition will be continued further. This study has acquainted student’s attitude, expectations and identified several barriers experienced in online classes by the students. Conformation of the medical education system with addressing the dilemmas can leads to a successful implementation of web based learning that will be most pertinent in this Covid 19 pandemic.

## Data Availability

All data are provided in manuscript

## Acknowledgement

Authors conveying heartiest thanks to Fahmina Tasmin Munia and Sadman Rafid for helping with the statistical suggestions.

